# Spatial clusters and social inequities in Covid-19 vaccine coverage among children in Brazil

**DOI:** 10.1101/2023.04.25.23289089

**Authors:** Alexandra Crispim Boing, Antonio Fernando Boing, Marcelo Eduardo Borges, Denis de Oliveira Rodrigues, Lorena Barberia, SV Subramanian

**Affiliations:** Federal University of Santa Catarina, Florianópolis, Brazil; Federal University of Goiás, Goiânia, Brazil; Sergio Arouca National School of Public Health, Oswaldo Cruz Foundation - Fiocruz, Rio de Janeiro, Rio de Janeiro, Brazil; University of Sao Paulo, Sao Paulo, Brazil; Department of Society, Human Development and Health, Harvard T. H. Chan School of Public Health, Boston, U.S.A.

**Keywords:** Covid-19, vaccination coverage, immunization, spatial analysis, children

## Abstract

This study examined the spatial distribution and social inequalities in COVID-19 vaccine coverage among children aged 5-11 in Brazil. First and second dose vaccine coverage was calculated for all Brazilian municipalities and analyzed by geographic region and deciles based on human development index (HDI-M) and expected years of schooling at 18 years of age. Multilevel models were used to determine the variance partition coefficient, and bivariate local Moran`s I statistic was used to assess spatial association. Results showed significant differences in vaccine coverage rates among Brazilian municipalities, with lower coverage in the North and Midwest regions. Municipalities with lower HDI and expected years of schooling had consistently lower vaccine coverage rates. Bivariate clustering analysis identified extensive concentrations of municipalities in the Northern and Northeastern regions with low vaccine coverage and low human development, while some clusters of municipalities in the Southeast and South regions with low coverage were located in areas with high HDI-M. These findings highlight the persistent municipal-level inequalities in vaccine coverage among children in Brazil and the need for targeted interventions to improve vaccine access and coverage in underserved areas.

## Introduction

The COVID-19 pandemic has brought unprecedented disruption to the lives of billions of people around the world. Since its emergence in 2019, it has caused around seven million deaths and remains a significant public health threat^1^. In addition to its high mortality rate, the virus has been linked to several negative health outcomes for survivors. These can include long-term respiratory, psychological, musculoskeletal, neurologic, and cardiovascular complications, which can have a profound impact on their quality of life^2^.

While the risk of severe illness and death from COVID-19 is higher among older adults and those with underlying health conditions, the disease has also had a serious impact on children. From March 2020 to March 3, 2023, more than 10,260 children between the ages of 5 and 11 have been hospitalized due to COVID-19 in Brazil. Among them, 2,514 required intensive care, and 467 deaths have been registered^3^.

The administration of vaccines, including for children, is critical in the fight against the COVID-19 pandemic. In Brazil, two vaccines have been approved for children aged 5 to 11 years old. In December 2021, the National Health Surveillance Agency (ANVISA) authorized the BNT162b2 vaccine, a mRNA vaccine developed by Pfizer-BioNTech, for use in this age group. The following month, authorization for the use of the CoronaVac vaccine, an inactivated-virus vaccine developed by Sinovac Life Sciences, was extended to children and adolescents between 6 and 17 years old who were not immunocompromised^4^. Despite extensive evidence demonstrating the safety and efficacy of vaccines against serious outcomes caused by COVID-19^5-7^, childhood vaccination in Brazil only began on January 14, 2022. The federal government held an unprecedented and heavily criticized public consultation to guide the Ministry of Health’s decision regarding the acquisition and purchase of pediatric vaccines. During this time and in the following months, intense campaigns of disinformation and discouragement of childhood vaccination took place in the country^8^.

Organized anti-vaccination movements, both inside and outside the country, have contributed to vaccine hesitancy, which, combined with other structural factors, has hindered broad vaccine coverage in Brazil. According to the World Bank, Brazil is one of the countries with the highest income inequality in the world^9^ and, despite having a public and universal health system, has a history of significant health disparities, including disparities in access to healthcare^10^. Furthermore, the country has recently experienced an economic crisis, reduced budgets for social programs, and adoption of austerity measures that have reduced public investment in health^11^.

International studies have identified inequalities in vaccination coverage against Covid-19 in children aged 5 to 11 years. In the United States (US), during the first three months of the vaccination campaign, significant racial, educational, and geographic disparities in vaccination coverage were observed ^12-14^. A recent study by Singh et al. (2022)^15^ that analyzed data from the first quarter of 2022 in the US also found a positive association between a child’s likelihood of being vaccinated and their parents’ education and income. In Brazil, a previous study has already identified significant area-level disparities in relation to covid-19 lethality and diagnostic testing performance^16^. It is crucial to extend the analysis of social inequalities in vaccination against covid-19 in Brazil, given the country’s history of social inequalities, high burden of covid-19, and mismanagement of the pandemic^17^.

The aim of the present study is to analyze the covid-19 vaccination coverage among children aged 5 to 11 years old in Brazil in 2022. The study investigated the association between vaccination coverage and municipal socioeconomic variables, and explored regional clusters of high or low coverage. Additionally, the study identified the geographic units that explain most of the outcome variability.

## Methods *Data sources Vaccine data*

We obtained the data from the National Immunization Program Information System (SI-PNI) available on the OPENDATASUS platform to determine the 2022 coverage of Covid-19 vaccinations in the 5 to 11-year-old population in the 5,570 Brazilian municipalities. The vaccine doses administered between epidemiological weeks 3 (beginning January 16, 2022) and 49 (beginning December 4, 2022) were included in our calculations. The SI-PNI is the official database of the Brazilian Ministry of Health that records all vaccine doses administered in the country, including Covid-19 vaccines, and is used to monitor vaccination coverage and support the development of immunization strategies. This database consists of anonymous records, including date and location of vaccine administration, vaccine type, dose type, and residency information of the vaccinated individuals by municipality and state.

Individuals that had more than six notifications in the database or lacked a unique identifier were excluded from the analysis. Records that lacked information on gender, vaccine administration date, municipality, or state were also removed. These excluded values are considered recording errors or incomplete records, and were therefore discarded to ensure the accuracy of our analysis.

To determine the vaccine dose (first or second), we utilized the order of administration instead of the date recorded by SI-PNI. The data was organized by epidemiological week, order of vaccine administration (dose one; dose two), municipality (n=5,570), state (n=26), and region (n=5). Using these groups, we calculated the cumulative number of doses administered to residents of each municipality, state, and region. The vaccine coverage data utilized in this study can be found at https://github.com/covid19br/dados-vacinas.

### Population data

The population data for children aged 5 to 11 years old by municipality, state, and region of residence was obtained from the Ministry of Health of Brazil (estimated in partnership with the Brazilian Institute of Geography and Statistics). To calculate the vaccine coverage, we divided the number of vaccinated individuals as recorded in SI-PNI by the resident population in this age group in each municipality, as provided by the Ministry of Health.

### Municipal socioeconomic data

The Municipal Human Development Index (HDI-M) and the expected years of schooling at 18 years were obtained for all 5,565 Brazilian municipalities in existence at the time of the 2010 National Census. The census was the primary source of socioeconomic data used in our study and was the most recent available. These indicators were calculated in collaboration with the United Nations Development Program, the Brazilian Institute of Applied Economic Research, and the José Pinheiro Foundation. The calculation of the HDI-M followed the same three dimensions of the global HDI - health, education, and income - and reflected local social progress and development (the higher the value, the greater the social development). However, it adapted the global methodology to the Brazilian context and the availability of national indicators. Further information can be found at http://www.atlasbrasil.org.br/. The expected years of schooling at 18 years of age refers to the average number of years of formal education that a cohort of children starting school is anticipated to complete by the time they reach 18 years of age, assuming that the current standards of education are upheld throughout their academic journey.

### Data analysis

Initially, we calculated the central tendency and dispersion measures of the coverage of the first and second doses of the vaccine. The statistics included the minimum and maximum values of vaccine coverage in Brazilian municipalities, the median, the 5th and 95th percentiles, and the standard deviation. Subsequently, the vaccine coverage was described for each region of Brazil, decile of the HDI-M, and decile of expected years of schooling at 18. In all cases, the estimates of the first dose application were calculated for 2022 for the months of March (two months after the start of the national vaccination campaign for children), July (six months after), and December (eleven months after). In the case of the second dose, the estimates presented were for the months of May (two months after the second dose was applied to the first children), September (six months after), and December (nine months after). To monitor the speed of vaccination, the vaccine coverage was also presented according to each of the epidemiological weeks included in the study. Vaccine coverage was limited to 100% of the population.

To assess the persistence of lower vaccination, we identified the 557 municipalities (10%) with the lowest coverage for the first dose in March 2022 and the 557 municipalities with the lowest coverage for the second dose in May 2022. We then checked whether these municipalities still had the lowest vaccination rates in the country after nine (first dose) to eleven months (second dose), or if they had moved to higher positions in terms of vaccination coverage.

Our data was organized into hierarchical groups, with municipalities as level 1, states as level 2, and regions as level 3. The Brazilian states and regions are presented in Supplementary Figure 1. We used multilevel linear models to determine the proportion of variation at each geographic level - municipalities, states, and regions. The variation in vaccine coverage (y) for municipality *i* in state *j* in region *k* was modeled as *y*_*ijk*_ = β_0_ + (ν_*0k*_ + υ_*0jk*_ + e_*0ijk*_), where ν_*0k*_ represents the random effect associated with regions, υ_*0jk*_ represents the random effect related to states, and e_*0ijk*_ represents the residual error for municipality i in state j. To determine the contribution of each level to the overall variation in vaccine coverage, we calculated the variance at each level (between-municipalities, between-states, and between-regions) and divided it by the total variance of the three levels. These analyses were performed using Stata 15.1 software.

Exploratory spatial analysis was performed to identify regional differences in first and second dose vaccination coverage in December 2022 across municipalities. We also performed spatial statistical analysis to detect spatial clusters of high and low vaccination coverage during the same period, based on the similarity of each municipality with its neighbors and the level of significance. We used the global Moran’s I statistic to assess the presence of spatial clustering in the dataset, and the local univariate Moran’s I statistic to identify clusters^18^. We classified municipalities with vaccination coverage similar to that of their neighbors and above the average as “high-high”, and those with coverage below the average but still similar to the neighborhood as “low-low”. Municipalities with vaccination coverage different from the neighborhood and with coverage both above and below the average were classified as “high-low” and “low-high”, respectively.

To assess the spatial association between vaccination coverage and the Municipal Human Development Index (HDI-M) and the expected years of schooling at 18, we used the bivariate local Moran’s I statistic^19^. These analyses capture the relationship between a municipality’s variable and the average of its neighbors for another variable. We classified municipalities with high and low vaccination coverage located in regions with socioeconomic indicators above or below the average as “high-high” and “low-low”, respectively. Municipalities with vaccination coverage different from the neighborhood, and with socioeconomic indicators above or below the average, were classified as “high-low” and “low-high”, respectively. Municipalities without a defined neighborhood were classified as “isolated”.

We evaluated the level of significance for the spatial association analyses with 999 simulations of the distribution by random permutation among the values, using a cutoff value of 0.05. Municipalities that fell outside the significance level were classified as “not significant”. All analyzed data is public and anonymized, and there was no need for ethical research committee approval.

## Results

During the analyzed period, 23,675,171 doses of the COVID-19 vaccines were administered to children aged 5 to 11 in Brazil, resulting in a vaccination coverage rate of 69.5% for the first dose and 46.1% for the second dose in December 2022. In the same month, half of the municipalities had population coverage equal to or greater than 75.0% of dose one and 53.4% of dose two. The regional distribution of vaccinations was significantly unequal among Brazilian municipalities, as shown by the dispersion measures presented in Table 1. Additionally, the observed standard deviation remained relatively unchanged from March to December 2022.

**Table 1.** Covid-19 vaccine coverage (%) in Brazilian municipalities among the pediatric population (5 to 11 years of age) by Municipal Human Development Index (HDI-M), expected years of schooling, and region of the country. Brazil, 2022.

|  | Dose 1 |  |  | Dose 2 |  |  |
| --- | --- | --- | --- | --- | --- | --- |
|  | March | July | December | May | September | December |
| Minimum | 2.4 | 3.8 | 4.2 | 0.4 | 1.1 | 1.2 |
| Maximum | 100.0 | 100.0 | 100.0 | 100.0 | 100.0 | 100.0 |
| Standard deviation | 20.5 | 20.4 | 20.1 | 20.5 | 21.6 | 21.9 |
| 5th percentile | 24.4 | 32.5 | 35.7 | 9.9 | 16.4 | 17.7 |
| Median | 60.8 | 71.4 | 75.0 | 40.7 | 51.4 | 53.4 |
| 95th percentile | 92.9 | 100.0 | 100.0 | 76.6 | 89.3 | 92.1 |
| **HDI-M (deciles)** |  |  |  |  |  |  |
| 1 (lowest) | 47.0 | 58.7 | 63.0 | 26.0 | 36.4 | 38.1 |
| 2 | 51.9 | 63.2 | 67.4 | 29.2 | 39.1 | 40.9 |
| 3 | 53.2 | 64.0 | 68.2 | 30.4 | 40.2 | 41.9 |
| 4 | 55.8 | 67.0 | 71.3 | 34.2 | 44.7 | 46.8 |
| 5 | 55.0 | 66.6 | 70.8 | 33.6 | 44.0 | 46.0 |
| 6 | 50.3 | 60.8 | 65.0 | 31.2 | 41.0 | 43.0 |
| 7 | 57.8 | 68.1 | 72.0 | 39.8 | 49.9 | 52.1 |
| 8 | 57.8 | 68.4 | 72.4 | 36.1 | 46.1 | 48.2 |
| 9 | 54.9 | 65.8 | 70.5 | 35.7 | 46.1 | 48.5 |
| 10 (highest) | 57.7 | 67.4 | 71.5 | 39.4 | 49.1 | 51.4 |
| **Expected years of schooling at 18 (deciles)** |  |  |  |  |  |  |
| 1 (lowest) | 40.2 | 51.4 | 55.3 | 22.7 | 31.7 | 33.4 |
| 2 | 49.0 | 59.9 | 64.1 | 29.3 | 39.4 | 41.3 |
| 3 | 50.2 | 61.5 | 65.6 | 29.7 | 39.6 | 41.4 |
| 4 | 50.2 | 61.3 | 65.5 | 28.8 | 38.9 | 40.6 |
| 5 | 54.1 | 65.4 | 69.8 | 31.4 | 41.9 | 43.9 |
| 6 | 53.5 | 65.9 | 69.4 | 31.4 | 41.7 | 43.7 |
| 7 | 56.2 | 66.9 | 71.0 | 34.1 | 43.9 | 45.9 |
| 8 | 59.5 | 69.8 | 74.1 | 40.7 | 51.0 | 53.4 |
| 9 | 60.4 | 70.6 | 74.7 | 41.8 | 52.1 | 54.5 |
| 10 (highest) | 62.0 | 71.6 | 75.7 | 43.4 | 53.6 | 56.1 |
| **Region** |  |  |  |  |  |  |
| North | 34.5 | 45.8 | 50.8 | 16.2 | 25.2 | 27.0 |
| Northeast | 55.7 | 66.8 | 71.0 | 32.6 | 42.9 | 44.7 |
| Southeast | 61.3 | 72.3 | 76.5 | 41.6 | 52.1 | 54.4 |
| South | 55.0 | 64.5 | 68.1 | 35.6 | 45.2 | 47.5 |
| Midwest | 40.3 | 50.7 | 55.1 | 20.7 | 30.7 | 32.5 |

Significant differences in vaccination coverage among Brazilian municipalities were observed when they were grouped according to their HDI-M and expected years of schooling at 18 years old. These differences existed in the application of both doses and throughout the analyzed period but were more pronounced in the second dose and the initial months of the onset of vaccination. Two months after the start of the vaccination campaign (March 2022), the decile with the highest HDI-M reached a coverage rate of 57.7% for the first dose, while the decile with the lowest HDI-M was at 47.0% (Table 1). The inequality was even greater when analyzing the two-month period after the start of the second dose campaign (May 2022), with coverage rates of 39.4% and 26.0% in the highest and lowest HDI-M deciles, respectively. By December 2022, vaccination coverage was still 13.5% higher in the municipalities with the highest HDI-M for the first dose and 34.9% higher for the second dose (8.5 and 13.3 percentage points (p.p.), respectively).

The differences were even more pronounced when the municipalities were grouped according to the expected years of schooling at 18 years old. When analyzing the application of the second dose we observed that the municipalities with the highest values of expected schooling had vaccination coverage 20.7 percentage points higher in May 2022 and 22.7 p.p. higher in December (Table 1). In all periods, the lowest vaccination coverage values were observed in the North and the highest in the Southeast region of the country. In the case of the second dose coverage, municipalities in the Southeast had a 156.8% higher coverage rate in May 2022 and 101.5% higher in December 2022.

The choropleth map of the first dose vaccine coverage showed higher coverage in the municipalities of the Southern, Southeastern, and Northeastern regions of the country, with a concentration of municipalities with low coverage in the Northern region and some parts of the Northeast and Midwest. Additionally, inter-municipal contrasts were observed within some states (Supplementary Figure 2). The second dose vaccine coverage map indicated that municipalities with rates below 50% were mainly concentrated in the Northern and Midwest regions, as well as in specific areas of the other regions (Supplementary Figure 2B). Univariate clustering analysis confirmed the concentration of low Covid-19 vaccine coverage (below average), with emphasis on the Northern, Midwest, and Northeast regions for both vaccine doses (Figure 1).

**Figure 1.**
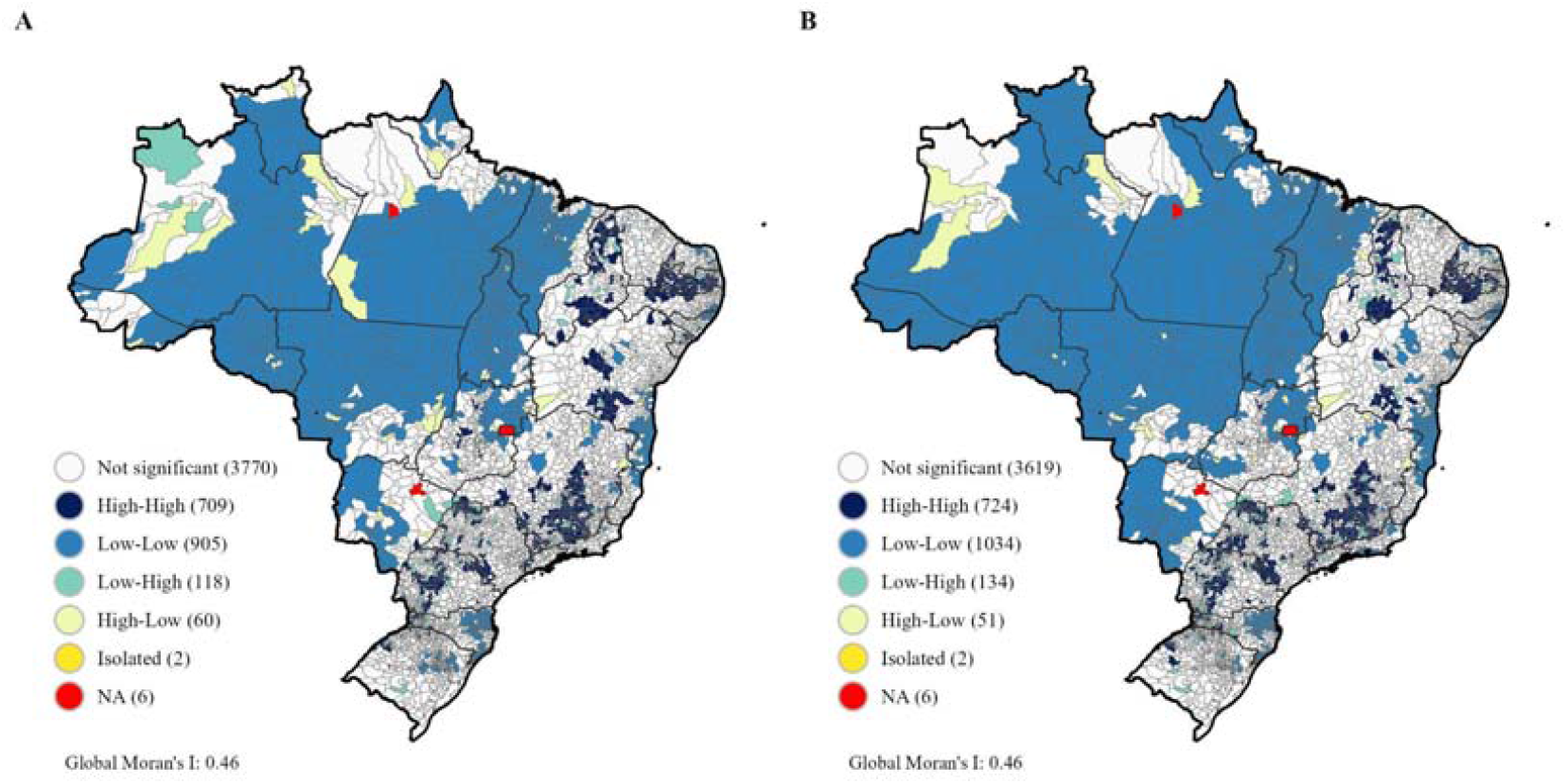
Univariate spatial association patterns in covid-19 vaccination coverage (%) of first dose (A) and second dose (B) clusters in Brazilian municipalities in the pediatric population (5 to 11 years old) in December 2022. Brazil, 2022.

The bivariate clustering maps revealed extensive concentrations of municipalities in the Northern and Northeastern regions that had low vaccine coverage and were located in areas of low HDI-M for both the first and second doses against Covid-19. In the Southeast and South regions, particularly in the states of São Paulo, Paraná, Santa Catarina, Rio Grande do Sul and Minas Gerais, some clusters of municipalities with low vaccine coverage were located in areas with high HDI-M (Figure 2A-2B). There were similarities in the analysis between vaccination coverage and expected years of schooling at 18 years of age, adding some states in the Midwest with low vaccination coverage and low expected years of schooling at 18 (Figure 2C-2D). In contrast, states such as Piauí, Paraíba, Ceará, Pernambuco and Bahia showed clusters of municipalities with high vaccine coverage located in areas of low HDI-M and low expected years of schooling at 18.

**Figure 2.**
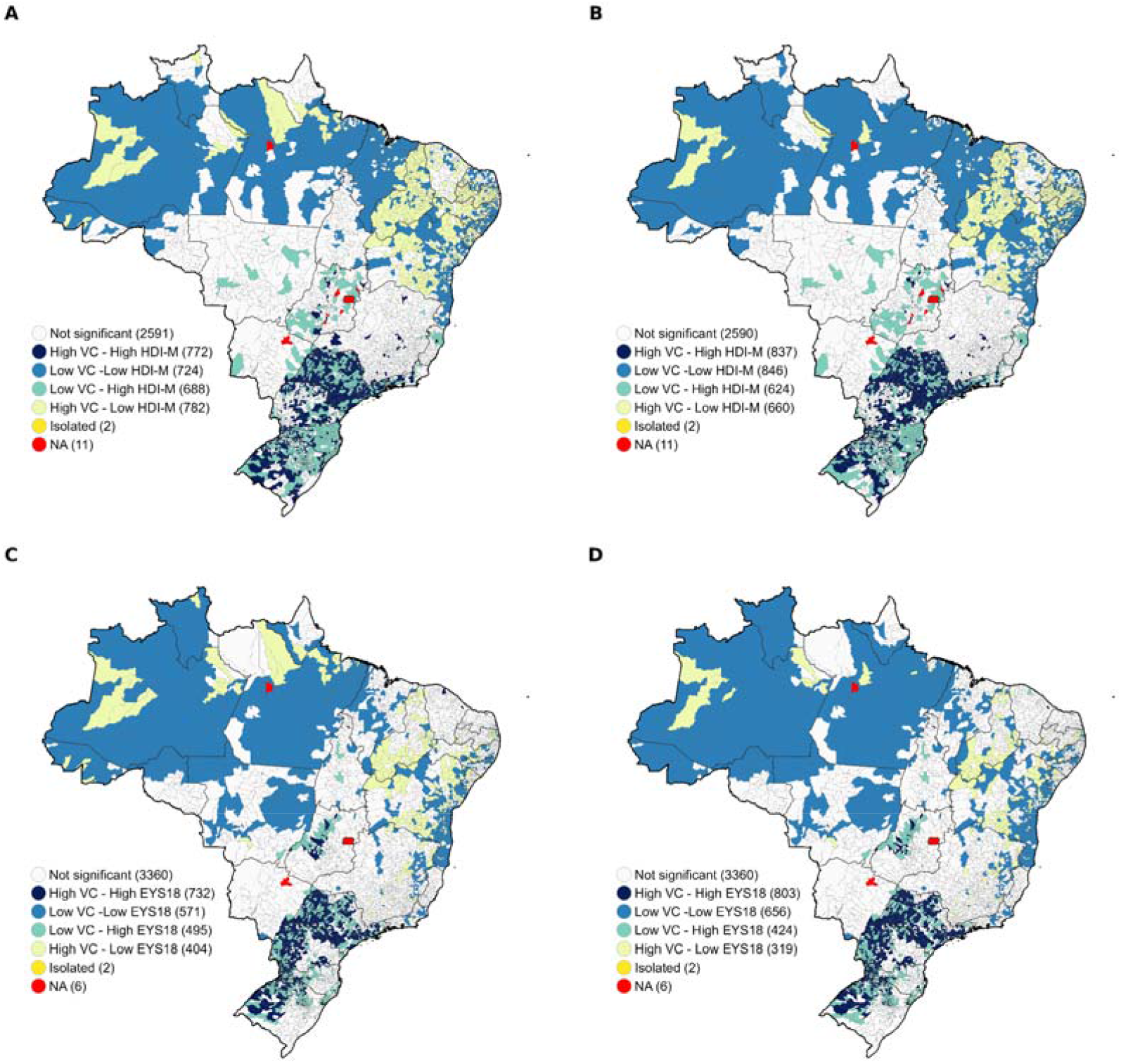
Patterns of bivariate spatial association between first dose covid-19 vaccination coverage (%) and HDI-M (A); second dose vaccination coverage (%) and HDI-M (B); first dose vaccination coverage (%) and Expected years of schooling at 18 (EYS18) (C); and second dose vaccination coverage (%) and Expected years of schooling at 18 (EYS18) (D) in Brazilian municipalities in the pediatric population (5 to 11 years old) in December 2022. Brazil, 2022.

The observed gradient of lower coverage among municipalities with the lowest HDI-M, lowest expected schooling at 18 years old, and located in the North and Midwest regions was evident in all the epidemiological weeks of 2022, as seen in Figure 3. In addition to the low vaccine coverage, particularly for the second dose, there is a distinct pattern of inequalities that remains consistent throughout the year.

**Figure 3.**
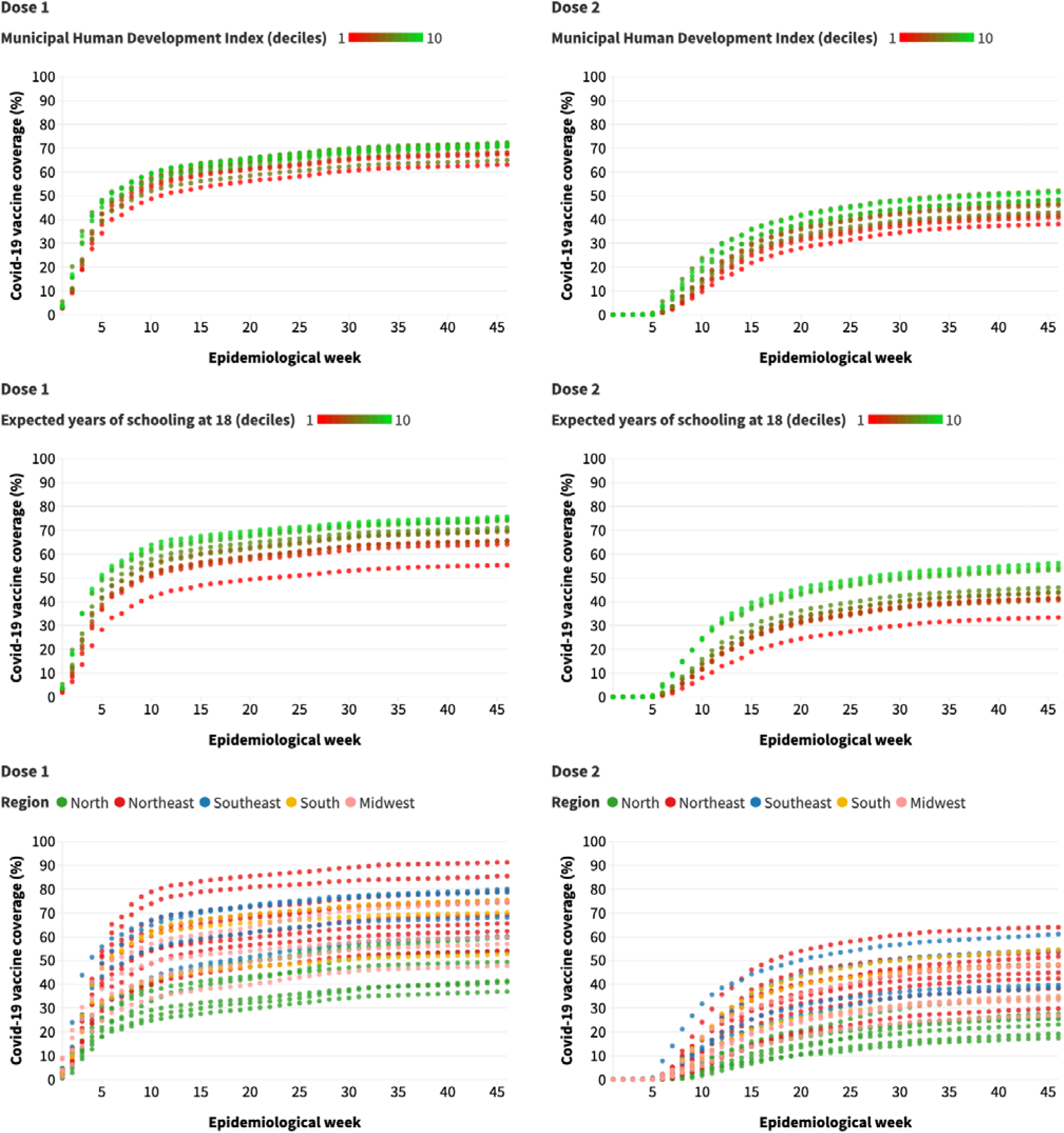
Covid-19 vaccine coverage (%) in Brazilian municipalities among the pediatric population (5 to 11 years old) according to the epidemiological week, Municipal Human Development Index (HDI-M), expected years of schooling at 18 years of age, and region of the country. Brazil, 2022.

The multilevel regression models showed that municipalities were the main source of variation in vaccine coverage. It was also observed that there were no important differences in the VPC estimates throughout the year or according to the dose administered. The between-municipality variation ranged from 59.5% (second dose, December 2022) to 67.7% (first dose, December 2022) of the total variation. Table 2 presents the variance estimates for each period and vaccine dose.

**Table 2.** Variance estimates, standard errors and variance partition coefficient (VPC) in vaccine coverage against Covid-19 in pediatric population (5-11 years-old). Brazil, 2022.

|  | Dose 1 | Dose 2 |
| --- | --- | --- |
| **May** |  |  |
| Region |  |  |
| VE (SE) | 118.35 | 116.90 |
| % VPC | 20.1% | 23.2% |
| State |  |  |
| VE (SE) | 103.93 | 87.42 |
| % VPC | 17.6% | 17.3% |
| Municipality |  |  |
| VE (SE) | 367.56 | 300.5 |
| % VPC | 62.3% | 59.5% |
| **September** |  |  |
| Region |  |  |
| VE (SE) | 100.84 | 120.81 |
| % VPC | 16.6% | 20.8% |
| State |  |  |
| VE (SE) | 98.71 | 92.36 |
| % VPC | 16.3% | 15.9% |
| Municipality |  |  |
| VE (SE) | 406.38 | 367.58 |
| % VPC | 67.1% | 63.3% |
| **December** |  |  |
| Region |  |  |
| VE (SE) | 98.01 | 120.37 |
| % VPC | 15.9% | 20.0% |
| State |  |  |
| VE (SE) | 100.37 | 94.26 |
| % VPC | 16.3% | 15.6% |
| Municipality |  |  |
| VE (SE) | 416.34 | 387.82 |
| % VPC | 67.7% | 64.4% |

VPC: variance partition coefficient; VE: variance estimate; SE: standard error

## Discussion

This study reports six main findings regarding childhood vaccination against COVID-19 in Brazil. Firstly, large regional disparities in vaccination coverage were observed during the first year of the vaccination campaign. Specifically, municipalities with the worst socioeconomic indicators had lower vaccination rates. Secondly, greater differences between municipalities were observed in the application of the second dose. Thirdly, worst vaccination coverage was mainly observed in the North and Midwest regions of the country. Fourthly, spatial analysis revealed municipal clusters demonstrating a correlation between immunization coverage and socioeconomic indicators. Fifthly, the municipalities with the lowest vaccination coverage at the beginning of the campaign remained mostly unchanged at the end of the year, indicating that these areas were not sufficiently prioritized and were left behind. Finally, multilevel analysis revealed that the majority of the variability in vaccination outcomes occurred at the municipal level.

Increasing vaccination coverage against Covid-19 for children aged 5 to 11 is crucial for public health. Although this age group is less susceptible to severe illness compared to adults and the elderly, children can still experience serious outcomes such as multisystem inflammatory syndrome, hospitalization, and even death^20^. Moreover, children contribute to community transmission of the virus^21^, making it essential that they are a priority in vaccination campaigns against Covid-19.

However, in several countries, vaccination coverage among children has been low. For example, in Italy, about four months after the start of the vaccination campaign, only 34% of children aged 5 to 11 had received their primary vaccine cycle^5,22^. In Australia, as of the end of September 2022, only 51.3% of children had received the first dose of the vaccine, and 40.4% had received the second dose^23^. In the United States, as of February 15, 2023, first dose coverage was 39%, and second dose coverage was 32% ^24^. In Brazil, as of December 2022 - eleven months after the start of the campaign, vaccination coverage was 69.5% and 46.1% for the first and second doses, respectively. In addition to the low national averages, our study identified that vaccination rates have been highly uneven within Brazil, which is consistent with what has been observed in other countries. In Australia, vaccination coverage varied substantially across the country, with differences of up to 400% between different locations ^23^. A similar pattern was seen in the United States, where lower vaccination coverage was observed in areas with greater vulnerability, particularly among black and Hispanic children compared to white children^12,13^.

Such inequalities emerge from multiple factors that involve different dimensions, such as the public policies implemented on the subject, the availability of health infrastructure, the logistical challenges faced^25^, and the economic and social conditions of the population^26^. Studies have shown that low income, lower educational levels, and geographic barriers affect access to vaccines^27^. In areas with greater poverty, there may be a lower supply of health services and greater difficulty in accessing them^28^, which can hinder the use of health services or force people to travel long distances in search of care^29,30^. In addition, lower family education and income have been associated with a lower likelihood of childhood immunization^31^ due to greater objective difficulties in prioritizing vaccination, accessing health services, or receiving adequate information on the importance of childhood vaccination within certain population groups^32^.

In the context of COVID-19 vaccination, a strong disinformation campaign has contributed to increased vaccine hesitancy among parents of children aged 5 to 11 years^33^. A study conducted in Canada found that a significant proportion of parents downplayed the risk of COVID-19 in their children^34^. Moreover, even among some parents who have been vaccinated against COVID-19 themselves, concerns about potential side effects have led to hesitancy in vaccinating their children. Doubts about vaccine safety and efficacy have also been identified as reasons for vaccine hesitancy^35^. Studies have shown that vaccine hesitancy is more common among less educated and Black parents^35^.

Our research also revealed lower COVID-19 immunization rates in the North and Midwest regions of Brazil. Historically, the North region has had the poorest indicators of population coverage of primary care, provision of doctors, nurses, and hospital beds in the country^36^. It also has the highest concentration, along with the Northeast, of states with poor social and economic indicators such as income, education, and poverty^37^. This combination of poor healthcare infrastructure and low socioeconomic indicators may have influenced the lower vaccination coverage observed in our study. Despite having some indicators of low social development, the Northeast region exhibited higher rates of child immunization against COVID-19. This may be attributed to the proactive role played by state governments in the region in formulating and executing policies to combat the pandemic^38^.

The present study has limitations that should be acknowledged. Firstly, the data on administered vaccine doses were obtained from the SI-PNI, which experienced problems in recording and transmitting data from municipalities during the pandemic, resulting in data lags and discrepancies. However, it is important to note that the SI-PNI is Brazil’s official system for registering immunizations and has been widely used for decades, so it is considered a reliable source of data despite the challenges encountered during the pandemic. A second limitation is that the municipal socioeconomic data used in the study were obtained from the 2010 census, which is now over a decade old. Additionally, the projections of resident population in each municipality during intercensal years, such as those included in this study, are based on the numbers obtained in the previous census. Therefore, there may be over or underestimation of residents in the municipalities, as well as changes in educational level and social development that have occurred since 2010. However, the grouping of municipalities into deciles in the analysis helped to minimize the impact of these potential fluctuations. In conclusion the inequalities in vaccination coverage should not be viewed as individual decisions isolated from the broader social, cultural, economic, and health determinants. In the case of COVID-19, factors such as delays in vaccine procurement, logistical challenges, lack of coordination, discouragement of childhood vaccinations, and childism have exacerbated existing inequalities.

To increase vaccination coverage in a country with significant social and health disparities, limited public budgets, and facing strong anti-vaccination campaigns, it is necessary to strengthen the public health system, expand social investments, and move beyond traditional vaccination campaigns. It is crucial to identify and acknowledge existing inequalities and barriers to healthcare access in order to implement policies and actions that can adapt services to address this reality. Information systems need to include data that allow people to track inequalities in real-time and make it easy to synthesize and publish information. Research needs to be conducted to better understand the reasons for vaccine hesitancy and the role of health services in low immunization rates. Primary care, healthcare professionals, and the community need to work together to implement strategies that are consistent with local realities. An intersectoral and transdisciplinary effort must be made to monitor and counter false, inaccurate, or out-of-context information, and robust and widespread campaigns to disseminate scientific information and combat misinformation are needed. Tackling inequalities cannot be a one-off effort and must be a top priority on the country’s political agenda. The pursuit of equity should be at the forefront of the design of all public policies to avoid promoting reverse equity.

## Data Availability

All data produced are available online at: https://github.com/covid19br/dados-vacinas

---

**Supplementary Figure 1.**
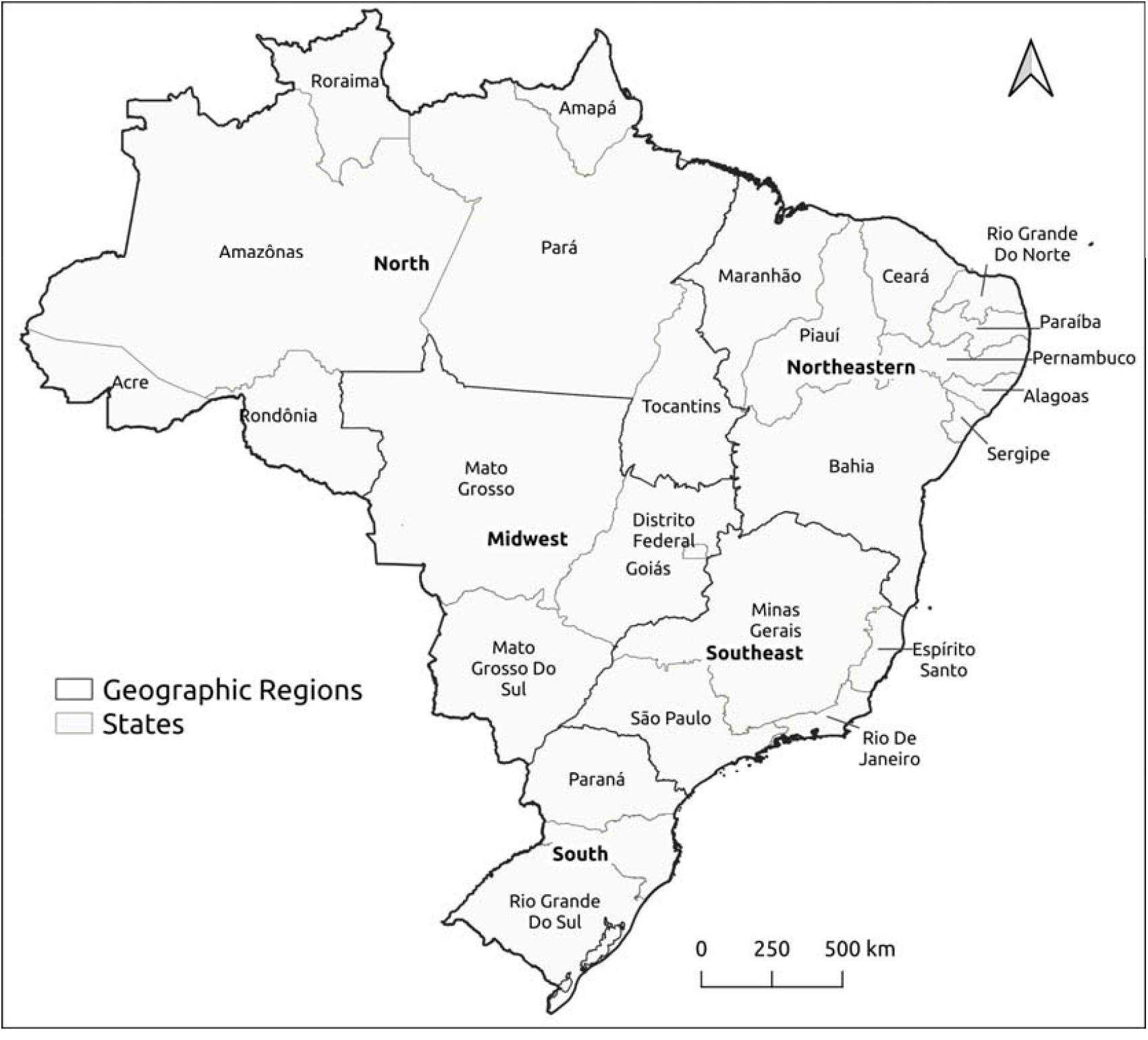
Brazilian states and regions.

**Supplementary Figure 2.**
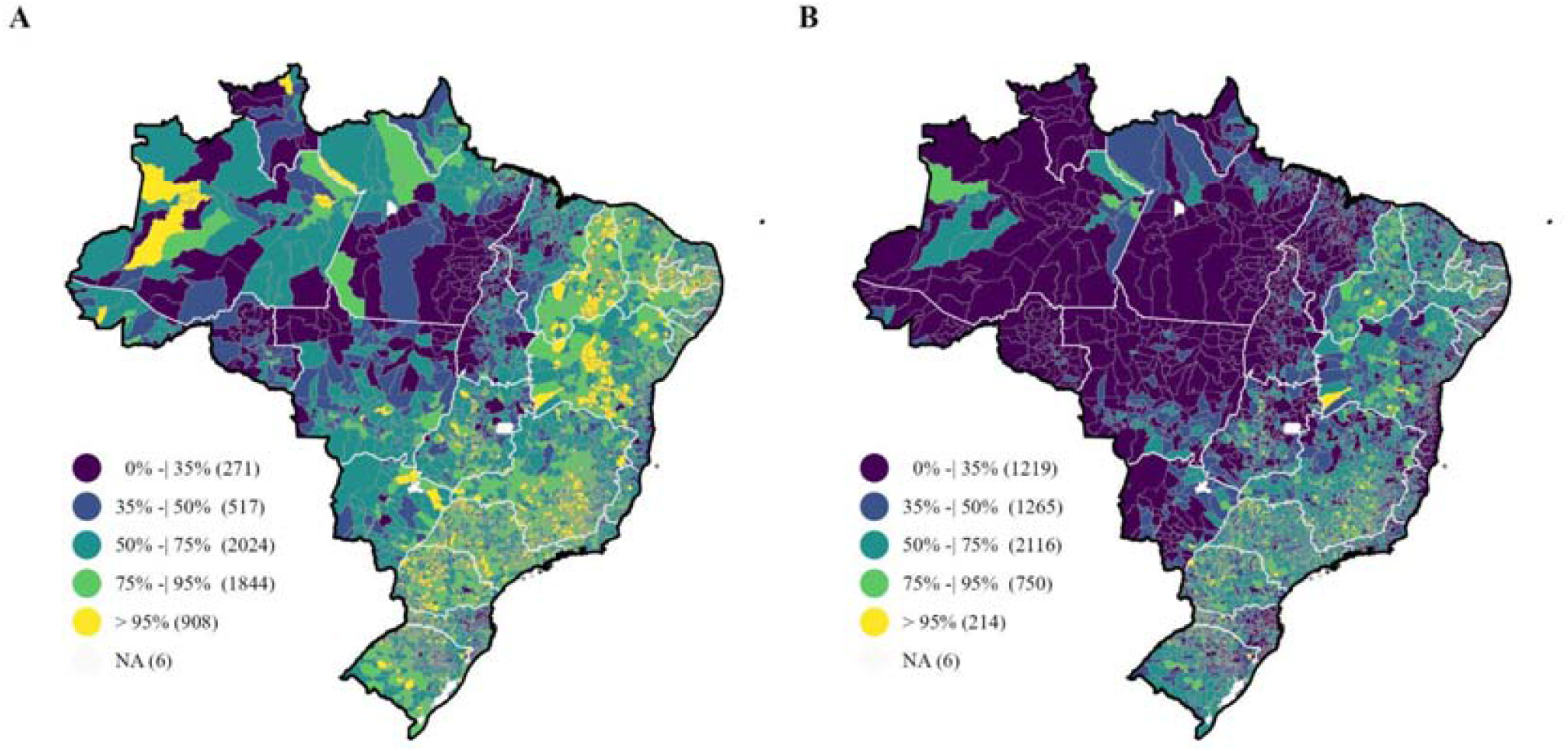
Covid-19 vaccination coverage (%) of the first dose (A) and second dose (B) in Brazilian municipalities among the pediatric population (5 to 11 years old) in December 2022. Brazil, 2022.

